# Association between epigenetic aging acceleration and amyloid biomarkers in bipolar disorder

**DOI:** 10.1101/2025.04.06.25325186

**Authors:** Gabriel R. Fries, Steven De La Garza, Ning O. Zhao, Andres W. Bass, Camila N. C. Lima, Nobuhide Kobori, Tatiana Barichello, Gustavo Turecki, Paul E. Schulz, Breno S. Diniz, Jair C. Soares

## Abstract

**Objectives:** Bipolar disorder (BD) has been associated with an elevated risk of Alzheimer’s Disease (AD). We assessed AD biomarkers in BD and tested whether epigenetic aging (EA) acceleration is a potential mechanism driving variability in these markers.

**Design, Setting, Participants:** Cross-sectional study of n=59 living individuals with BD and n=20 age- and sex-equated control participants, as well as analyses of postmortem brain samples (Brodmann area 9/46) from n=46 individuals with BD.

**Measurements:** Amyloid beta (Aβ)_40_, Aβ_42_, and total Tau levels were measured in plasma from individuals with BD and controls, and Aβ_42_ levels were measured in brains. EA and its acceleration (blood: GrimAge and DunedinPACE; brains: DNAmClock_Cortical_) were estimated for all samples. Individuals with BD were split into quartiles with accelerated or slower EA if they were in the first or fourth quartiles for GrimAge acceleration (AgeAccelGrim), DunedinPACE, or DNAmClock_Cortical_ acceleration (DNAmClock_Cortical_Accel).

**Results:** Individuals with BD showed an increase in Aβ_40_ (p=.049) and a decrease in the Aβ_42/40_ ratio (p=.035) compared to controls. A decrease in the Aβ_42/40_ ratio was also found in individuals with BD with high versus low AgeAccelGrim (p=.028). Brain Aβ_42_ levels significantly correlated with DNAmClock_Cortical_Accel (r^2^=.270, p=.007), with those with high EA acceleration showing higher brain Aβ_42_ after controlling for confounders (p=.008).

**Conclusions:** Our results provide preliminary evidence that EA may explain the variability in AD risk in individuals with BD and could act as a target for preventing dementia and AD in BD.

## 1. Introduction

Bipolar disorder (BD), a recurring and often debilitating psychiatric disorder with a prevalence of around 2%, has been associated with premature aging features and abnormalities in biological aging processes.^1^ These phenomena are manifested by age-related physiological changes,^2^ accelerated brain aging,^3^ and increased morbidity and premature mortality.^4^ A faster age-related decline in executive function and global cognition in BD has also been reported,^5^ although recent longitudinal studies have not supported these initial findings.^6^ While the molecular mechanisms underlying the premature aging phenotype in BD are largely unknown, it is hypothesized to involve biological pathways related to accelerated biological aging.

Epidemiological, pathophysiological, and clinical data suggest a strong relationship between BD and Alzheimer’s disease (AD),^7^ with individuals with BD showing an increased risk of developing AD^8–11^ (hazard ratio = 2.37^10^ – 10.37).^9^ Both conditions share a robust polygenic overlap,^12^ and some individuals with BD develop neurodegenerative alterations resembling those observed in AD.^13^ BD has been associated with a reduction in gray matter volumes, which emerges primarily in older patients compared to age-matched controls.^14^ Other studies also found decreased concentrations of the soluble forms of amyloid precursor protein (sAPP)α and sAPPβ, differences in the ratios of amyloid β (Aβ)_42/40_ and Aβ_42/38_ in individuals with BD,^15^ increased plasma levels of glial fibrillary acidic protein and neurofilament light chain,^16^ and an association between cerebrospinal fluid (CSF) ratios of Aβ_42/40_ and Aβ_42/38_ and altered cognitive performance in BD.^17^ Notably, investigations exploring neurodegeneration in BD have not always found significant and replicable results,^18^ suggesting variability in neurodegenerative processes in this population. This inconsistency parallels the heterogeneity observed in clinical features and cognitive performance in BD^19^ and supports the need for more accurate ways of identifying individuals with BD at high risk of neurodegenerative processes and cognitive decline.

Accelerated biological aging, as measured by epigenetic aging (EA) biomarkers, has been consistently linked to BD^20,21^ and could be the missing link underlying the observed variability in neurodegeneration and premature aging phenotypes in BD. The goal of this study was to assess biomarkers of neurodegeneration and AD in both blood and postmortem brains of individuals with BD to test whether EA acceleration is a potential mechanism driving variability of dementia and AD risk in BD. Our primary hypothesis is that changes in biomarkers of neurodegeneration and AD are primarily found in individuals with BD presenting with accelerated EA.

## 2. Methods

### Participants

We included N = 59 individuals with a diagnosis of BD type I (BD-I) and N = 20 age- and sex-equated non-psychiatric controls recruited at the Center of Excellence in Mood Disorders at The University of Texas Health Science Center at Houston (**Table 1**). The BD-I diagnosis was ascertained with the Structured Clinical Interview for DSM-IV Axis I Disorders (SCID-I). Interviews were administered by trained evaluators and reviewed by a board-certified psychiatrist. Acute manic and depressive symptoms were assessed by the Young Mania Rating Scale (YMRS) and Montgomery-Asberg Depression Rating Scale (MADRS), respectively.

**Table 1.**
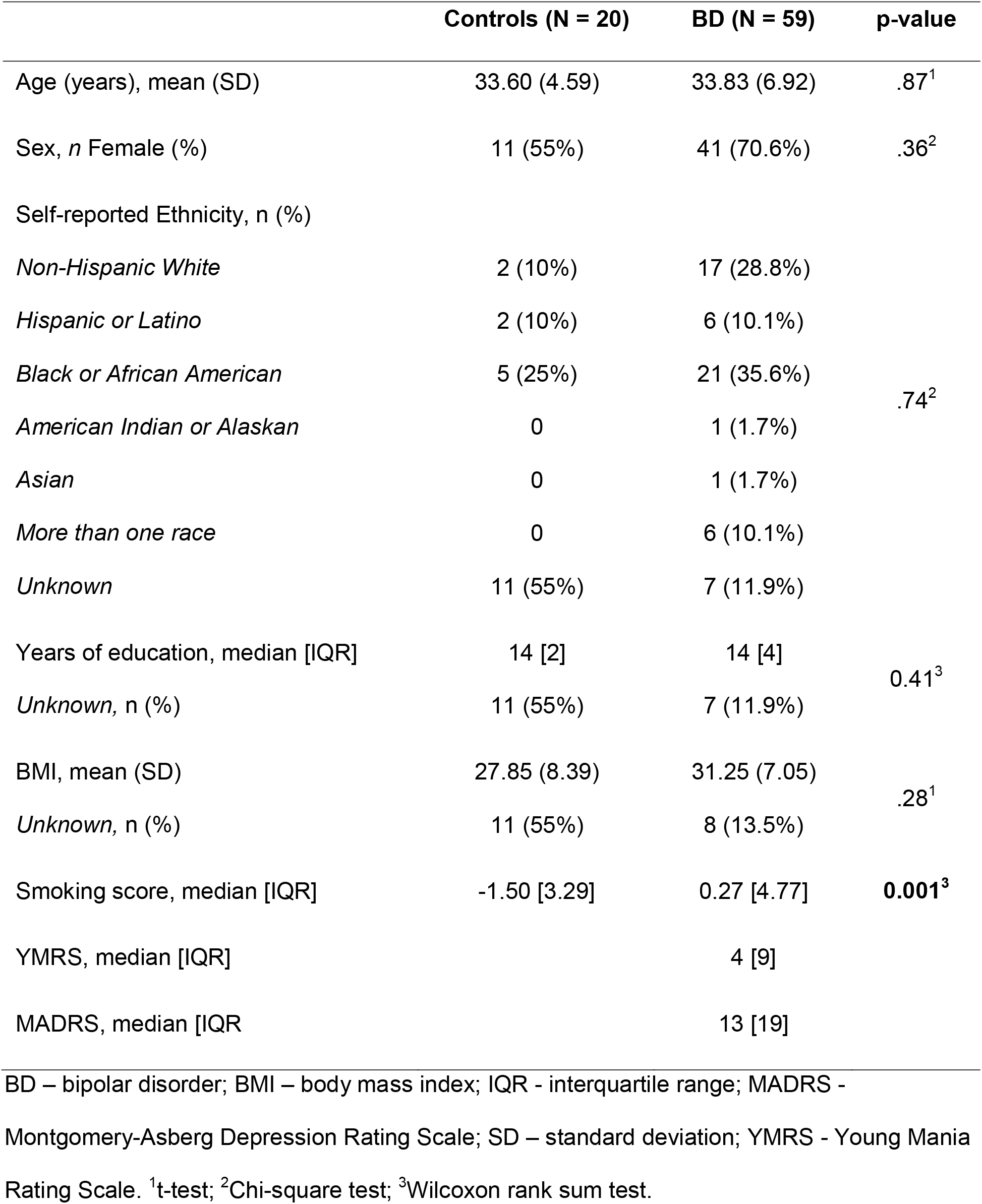
Sample demographics.

Control individuals did not have a history of BD or any lifetime diagnosis of psychiatric disorders. Exclusion criteria for all participants (BD and controls) included any neurological disorders and traumatic brain injury, schizophrenia, developmental disorders, intellectual disability, and recent illicit drug use by urine drug screen. Exclusion criteria for the non-psychiatric control participants also included a history of any Axis I disorder in first-degree relatives or having taken a prescribed psychotropic medication at any point in their lives. The study protocol was approved by the local institutional review board (IRB), and informed consent was obtained from all participants at enrolment and prior to any procedure.

### Postmortem brains

Postmortem brain samples (Brodmann area (BA) 9/46 of the dorsolateral prefrontal cortex) from N = 46 individuals with a diagnosis of BD (24 female, mean ± standard deviation age 66.16 ± 8.76) were obtained through the NIH NeuroBioBank (including samples from The Human Brain and Spinal Fluid Resource Center, Harvard Brain Tissue Resource Center, National Institute of Mental Health Human Brain Collection Core, University of Maryland Brain and Tissue Bank, and the University of Miami Brain Endowment Bank), and the Douglas-Bell Canada Brain Bank. The diagnosis was confirmed through a psychological autopsy with the next-of-kin, and information on the postmortem interval (PMI), age at death, sex, race (Caucasians), comorbidities, manner of death (accidental, natural, or suicide), and neuropathological findings were recorded for all subjects (**Supplementary Table 1**).

### AD biomarkers

Peripheral blood was collected from all living participants into EDTA-containing vacutainers, followed by separation of plasma and buffy coat and storage at −80^°^C until downstream analyses. Plasma levels of Aβ_40_, Aβ_42_, and total Tau were measured in duplicate in all samples with the Simoa® Neurology 3-Plex A Advantage Kit for SR-X (Quanterix). We also assessed Aβ_42_ levels in the postmortem brain samples using the Human Amyloid Beta 42 ELISA Kit (ab289832, Abcam), following the manufacturer’s instructions for tissue homogenate processing and assaying. Of note, levels of Aβ_40_ were not measured in the brain samples due to limited sample availability.

### DNA methylation and EA estimates

DNA was isolated from buffy coat samples using the DNeasy Blood & Tissue Kit (Qiagen) and from the postmortem brain samples using the *Quick*-DNA/RNA™ Miniprep Plus Kit (Zymo Research), followed by quantification on NanoDrop (Thermo Fisher). Five hundred nanograms of DNA were bisulfite-converted with the EZ DNA Methylation Kit (Zymo Research), followed by the assessment of genome-wide DNA methylation levels with the Infinium EPICBeadChip v1.0 (Illumina) in an iScan microarray scanner (Illumina), according to the manufacturer’s instructions. Measures of EA (GrimAge^22^ and DunedinPACE^23^ in blood and DNAmClock_Cortical_^24^ in brain) were estimated with the DNA Methylation Age Calculator (dnamage.genetics.ucla.edu/, GrimAge) and the R packages *DunedinPACE* and *dnaMethyAge*^*23,25*^ for DunedinPACE and DNAmClock_Cortical_, respectively. EA acceleration was obtained by regressing the estimated GrimAge or DNAmClock_Cortical_ on chronological age, using the residuals as aging acceleration indices (AgeAccelGrim or DNAmClock_Cortical_Accel). The proportion of neurons in the postmortem brain samples was estimated from the DNA methylation data with the R package *CETS*.^*26*^ Finally, a ‘smoking score’ reflecting smoking behavior and long-term exposure was estimated in blood samples with the R package *EpiSmokEr*^*27*^, providing increased accuracy over self-reported smoking information. Since the smoking score was developed for use exclusively in blood, we took the methylation M-values levels of the CpG cg05575921 as a reliable proxy of smoking in the postmortem brain samples, as previously suggested^28^.

### Statistical analyses

The Shapiro-Wilk test was used on all quantitative variables to test for normality, followed by group comparisons with the Welch Two Sample t-test (parametric variables), Wilcoxon rank sum test (non-parametric variables), Pearson’s Chi-squared test (categorical variables), and with generalized linear models controlling for age and sex. Individuals with BD were further split into quartiles based on the DunedinPACE variable and AgeAccelGrim variable to explore the association between AD biomarkers and accelerated aging. Generalized linear models were also used to assess the association between z-scores of DNAmClock_Cortical_Accel and Aβ_42_ levels measured in the brains of those diagnosed with BD with or without controlling for age, sex, and PMI. Finally, brain samples were split into quartiles using the DNAmClock_Cortical_Accel variable, followed by a linear model comparing Aβ_42_ levels between the first and fourth quartiles while controlling for age, sex, PMI, neuronal proportion, and smoking. Two-sided P-values<.05 were taken as statistically significant.

## 3. Results

There were no statistically significant differences between individuals with BD and controls in chronological age, sex, race/ethnicity, years of education, or body mass index (BMI) (**Table 1**). The majority of individuals with BD included were medicated (86.4%), presenting with mild symptoms of depression (median [interquartile range (IQR)] MADRS −13 [19]) and no symptoms of acute mania (median [IQR] YMRS −4 [9]). Details on medication use and psychiatric comorbidities in the individuals with BD are shown in **Supplementary Table 2**.

We found no statistically significant differences between individuals with BD and control participants for plasma Aβ_42_ or total Tau levels (p > .05, **Table 2**). Individuals with BD, however, showed a significant increase in Aβ_40_ (p = .049) and a decrease in the Aβ_42/40_ ratio compared to controls (p = .035), which remained statistically significant after controlling for age and sex (p =.044 for Aβ_40_ and p = .009 for Aβ_42/40_). Individuals with BD also showed a higher AgeAccelGrim (p <. 001) and DunedinPACE (p = .002) compared to control participants (**Table 2**), in line with previous studies showing a significant EA acceleration in BD.^20,21^ While DunedinPACE differences between BD and controls remained statistically significant after controlling for chronological age, sex, and smoking scores (β = −.09, p = .009), that was not the case for AgeAccelGrim (β = −.92, p = .327). Finally, we found a negative correlation between the Aβ_42/40_ ratio and DunedinPACE in the whole sample (N = 79, r^2^ = −.2, p = .008, **Supplementary Figure 1 and Supplementary Table 3**). No other significant association was found between AD biomarkers and estimates of accelerated EA (AgeAccelGrim or DunedinPACE), either in the whole sample or when assessing the associations separately within individuals with BD and controls (p > .05 for all, **Supplementary Table 3**).

**Table 2.** Alzheimer’s disease and epigenetic aging biomarkers in individuals with bipolar disorder and controls.

| Predictor | Control<br>(N = 20) | BD<br>(N = 59) | p-value |
| --- | --- | --- | --- |
| Aβ<sub>40</sub> (pg/mL), mean<br>(SD) | 162.59<br>(40.07) | 183.85<br>(40.32) | **.049<sup>1</sup>** |
| Aβ<sub>42</sub> (pg/mL), mean<br>(SD) | 9.74<br>(1.37) | 9.94<br>(1.89) | .61<sup>1</sup> |
| Total Tau (pg/mL),<br>mean (SD) | 4.03<br>(1.35) | 3.75<br>(1.23) | .41<sup>1</sup> |
| Aβ<sub>42/40</sub> ratio, median<br>[IQR] | 0.058<br>[0.008] | 0.056<br>[0.006] | **.035<sup>2</sup>** |
| AgeAccelGrim, mean<br>(SD) | -1.35<br>(3.38) | 1.78<br>(4.50) | **<.001<sup>1</sup>** |
| DunedinPACE, mean<br>(SD) | 0.93<br>(0.14) | 1.07<br>(0.14) | **.002<sup>1</sup>** |

Aβ – amyloid beta; BD – bipolar disorder; IQR – interquartile range; SD -standard deviation. <sup>1</sup>t-test, <sup>2</sup>Wilcoxon Rank-Sum Test.

When comparing individuals with BD with accelerated or slower EA based on quartiles, we found a significant decrease in the Aβ_42/40_ ratio in those with high AgeAccelGrim (p = .028, **Table 3**), although this difference did not remain significant after controlling for age, sex, and smoking score (p = .223). No differences were found in any AD biomarker between subgroups of individuals with BD based on DunedinPACE (**Supplementary Table 4**).

**Table 3.** Analysis of differences in quartiles of the AgeAccelGrim variable (blood samples)

| Predictor | 1<sup>st</sup> quartile<br>(N = 15) | 4<sup>th</sup> quartile<br>(N = 15) | p-value |
| --- | --- | --- | --- |
| Age, mean (SD) | 35.47 (8.18) | 33.47 (5.84) | .45<sup>1</sup> |
| Sex, *n* Female (%) | 11 (73%) | 7 (47%) | .26<sup>2</sup> |
| BMI, mean (SD) | 30.59 (8.19) | 28.67 (6.66) | .54<sup>1</sup> |
| Smoking score, median<br>[IQR] | -1.76 [1.72] | 5.20 [8.20] | **<0.001**<sup>3</sup> |
| Aβ<sub>40</sub> (pg/mL), mean (SD) | 187.85<br>(38.66) | 194.27 (41.17) | .66<sup>1</sup> |
| Aβ<sub>42</sub> (pg/mL), mean (SD) | 10.65 (2.14) | 10.16 (1.88) | .51<sup>1</sup> |
| Total Tau (pg/mL), mean<br>(SD) | 3.81 (.78) | 3.72 (1.46) | .084<sup>1</sup> |
| Aβ<sub>42/40</sub>, median [IQR] | .057 [.002] | .052 [.006] | **.028**<sup>3</sup> |
| AgeAccelGrim, median<br>[IQR] | -3.13 [2.71] | 7.39 [3.16] | **<.001**<sup>3</sup> |
| DunedinPACE, mean<br>(SD) | 0.96 (.14) | 1.14 (.12) | **<.001**<sup>1</sup> |

Aβ – amyloid beta; BD – bipolar disorder; BMI – body mass index; IQR – interquartile range; SD – standard deviation. <sup>1</sup>t-test; <sup>2</sup>Chi-square test; <sup>3</sup>Wilcoxon rank sum test.

While blood-based biomarkers are crucial in the clinical setting, a direct measure of both EA and AD alterations in brain tissues is key to exploring pathophysiological mechanisms underlying this association. In this context, brain-specific EA markers have been recently developed, among which the DNAmClock_Cortical_ stands out given its intended focus on improving the accuracy of age prediction specifically in human cortex tissue.^24^ We found a significant association between Aβ_42_ levels and DNAmClock_Cortical_Accel in postmortem prefrontal cortex samples of individuals with BD (Spearman’s rho = .304, p = .039, **Fig. 1**). This effect remained significant after controlling for age, sex, and PMI (adjusted r^2^ = .270, p = .007). Finally, similar to our analyses in blood, we further split the postmortem brain samples into quartiles based on the DNAmClock_Cortical_Accel variable and compared the Aβ_42_ levels between the first (slowest EA) and fourth (highest EA acceleration) quartiles. Brain quartile groups did not differ for confirmed AD-related neuropathological findings or frequency of suicide death (p > .05, **Table 4**). As shown in **Table 4**, individuals with BD with the highest EA acceleration showed significantly higher brain Aβ_42_ levels than did those from the first quartile (p = .020), which remained significant even after controlling for age, sex, PMI, neuronal proportion, and methylation levels (M-values) of cg05575921 (proxy of smoking in postmortem brains,^28^ p = .008).

**Figure 1.**
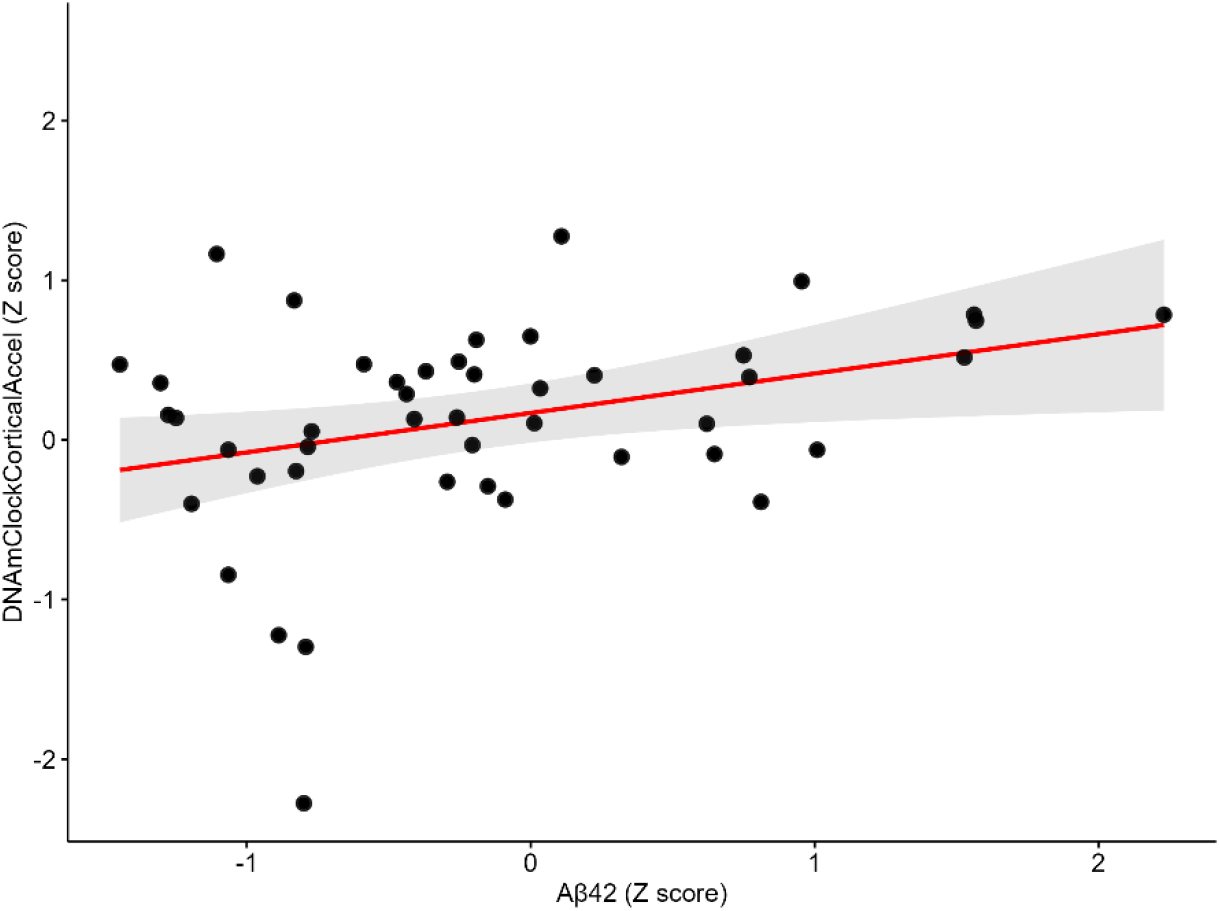
Association between DNAmClock_Cortical_Accel (Z-scored) and amyloid beta (Aβ)_42_ levels (Z-scored) in N = 46 postmortem brain samples (prefrontal cortex, Brodmann area 9/46) of individuals with bipolar disorder. General linear model controlled for age, sex, and postmortem interval (adjusted r^2^ = .270, p = .007)

**Table 4.** Analysis of differences in quartiles of the DNAmClock_Cortical_Accel variable (postmortem brain samples)

| Predictor | 1<sup>st</sup> quartile<br>(N = 12) | 4<sup>th</sup> quartile<br>(N = 12) | p-value |
| --- | --- | --- | --- |
| Age, mean (SD) | 66.64<br>(11.03) | 64.28<br>(7.99) | 0.600<sup>1</sup> |
| Sex, *n* Female (%) | 3 (25%) | 9 (75%) | **0.041**<sup>2</sup> |
| PMI, mean (SD) | 24.83<br>(13.10) | 34.67<br>(12.61) | 0.074<sup>1</sup> |
| Aβ<sub>42</sub> (z-score), median [IQR] | -0.80 [0.77] | 0.43 [1.74] | **0.020**<sup>3</sup> |
| DNAmClock<sub>Cortical</sub>Accel (z-score), median [IQR] | -0.39 [0.96] | 0.77 [0.30] | **<0.001**<sup>3</sup> |
| Neuronal proportion, mean (SD) | 0.38 (0.15) | 0.26 (0.14) | 0.050<sup>1</sup> |
| Smoking proxy (cg05575921)<br><sup>28</sup>, M-values, mean (SD) | 0.20 (1.00) | -0.08<br>(1.03) | 0.500<sup>1</sup> |
| Confirmed AD neuropathology,<br>*n* (%) | 1 (8.3%) | 1 (8.3%) | 1.000<sup>2</sup> |
| Suicide death, *n* (%) | 3 (25%) | 6 (50%) | 0.206<sup>2</sup> |

Aβ – amyloid beta; AD - Alzheimer's diseases; BD – bipolar disorder; PMI - postmortem interval; SD – standard deviation. <sup>1</sup>Welch Two Sample t-test; <sup>2</sup>Pearson's Chi-squared test; <sup>3</sup>Wilcoxon rank sum exact test.

## 4. Conclusions

BD has been linked to cognitive impairment and a higher risk of dementia and AD; hence, identifying the mechanisms underlying these associations is essential to developing novel interventions to mitigate or prevent the onset of these conditions in high-risk individuals. Our results suggest that EA may be an important source of variability in AD biomarkers in both blood and postmortem brains, where significant changes in AD-related biomarkers are primarily detected in individuals with BD presenting with accelerated EA. To our knowledge, this is the first study to show an increased pace of epigenetic aging (DunedinPACE) in BD and to explore the link between biomarkers of AD and biological aging measures in BD. Our exploratory findings may pave the way for future studies specifically targeting individuals with BD who show accelerated EA and may benefit from strategies to prevent the onset of dementia and AD.

The link between BD and AD has been consistently reported in the literature. A recent study in the UK BioBank, for example, demonstrated that a history of BD is associated with a significant increase in the risk of developing AD, even after adjusting for many potential confounders such as age, sex, educational level, diabetes, BMI, smoking, and the *APOE* ε4 allele.^10^ The same study also found a nominally significant causal association between BD and AD and a significant (false discovery rate-corrected) association between BD and a family history of AD tested with Mendelian randomization methods.^10^ Such a causal association between BD and AD was also recently reported in an independent sample,^8^ which is in line with previous studies reporting an increased risk of AD^9^ and dementia^11^ in individuals with BD.

We found a significant increase in the plasma Aβ_40_ and a decrease in the plasma Aβ_42/40_ ratio in individuals with BD compared to controls, which are suggestive of abnormal Aβ processing in the brain associated with AD pathology. Importantly, the plasma Aβ_42/40_ ratio has been shown to have a stronger correlation with brain Aβ burden and a better diagnostic and prediction accuracy than Aβ_42_ or Aβ_40_ alone, in addition to correcting for pre-analytical and analytical confounders.^29^ In addition, lower plasma Aβ_42/40_ has been associated with greater cognitive decline,^30^ brain Aβ burden assessed by positron emission tomography (PET),^31^ and CSF Aβ_42/40_ levels,^32^ and has been used to detect older adults at the early stages of AD or at higher risk of developing dementia upon follow-up.^29,32^ Therefore, in tandem with the current literature, our findings provide a mechanistic link that supports the epidemiological association between BD and AD.

Of note, not all studies have reported a significant association between AD biomarkers and BD.^18^ While this has at times been interpreted as a lack of evidence linking these conditions, it may also be explained by a significant variability in the risk of neurodegenerative changes and AD presented by individuals with BD and the heterogeneity of biological mechanisms of both conditions. Our findings showing EA in BD and its association with brain amyloid biomarkers suggest that EA may help identify more homogeneous subgroups of individuals with BD who are at higher risk for dementia and AD. Consistent with this, a previous study from our group found a negative association between EA acceleration and cognitive function in adults with BD,^21^ a finding supported by reports showing an association between accelerated EA and impaired cognitive performance.^33^ Studies have also shown significant associations between accelerated EA and AD and its neuropathological markers,^34^ although not consistently across all datasets,^35^ further supporting the hypothesis of a link between EA acceleration, cognitive decline, and risk of dementia and AD in BD. Our findings are even more remarkable since our cohort is very young (median age of 34 years), suggesting that EA acceleration and neurodegenerative changes in the brain already take place much earlier than the expected Aβ build-up in the brain of individuals with late-onset AD. Overall, our results offer evidence for potential mechanisms supporting the hypothesis that BD may be a neuroprogressive disorder.

Importantly, differences in plasma Aβ_42/40_ ratio between groups did not remain significant after controlling for smoking, suggesting it as a potential mechanism driving the EA and its effects in the context of BD. Indeed, smoking scores were higher in our cohort of individuals with BD compared to control participants. In the same vein, smoking has been associated with accelerated EA in different populations, including in individuals with BD,^21^ as well as with an increased risk of dementia and AD.^36^ While this suggests that smoking may be a driver of EA and alterations in AD biomarkers in the context of BD, we did not find a significant effect of smoking in the association between EA and Aβ_42_ levels in the postmortem brain analyses. While preliminary, this finding supports a possible link between EA acceleration and AD risk in BD that may not be fully accounted for by the effects of smoking, warranting future studies inquiring about independent mechanisms.

Our results should be viewed in light of the study’s many limitations. Firstly, the analyses were performed on a small sample size, particularly after splitting the sample into smaller groups based on quartiles. For this reason, and although we attempted to control our analyses for many potential confounders, not only do we acknowledge the potential for false positives, but our results also need to be taken as exploratory and in need of further replication in larger sample sizes. There are also inherent limitations associated with the assessment of neurodegenerative and AD-related biomarkers in plasma when compared to CSF. Moreover, the cross-sectional design and the inclusion of young individuals prevent causal interpretation and proper inferences regarding long-term aging acceleration and neurodegeneration in BD. Conversely, our results suggest that EA, and possibly neurodegeneration, is an early event in BD that can contribute to the neuroprogressive changes commonly observed in these individuals.

Interestingly, positive amyloid PET scans can be found up to twenty years before the onset of AD,^37^ as is the case for plasma levels of measures related to neurodegeneration.^38^ Future studies should focus on mid-to late-life individuals to adequately test these associations.

Additional limitations that we could not directly address include the effects of medications, which may act as confounders in the analyses (especially in light of the literature showing an important aging-modulating effect of medications used for BD treatment and the reported protective effect of lithium on neurodegenerative processes),^39^ as well as a lack of assessment of other variables known to affect AD risk, such as hypertension^40^ and *APOE* ε4 genotype. Accordingly, there was also limited assessment of AD biomarkers in our postmortem brains (e.g., Aβ_40_, neuritic plaques, Tau), as well as potential confounding effects of comorbidities in both the living cohort and the postmortem brains. Specifically in the latter, some individuals included in our analysis already showed AD-related neuropathological differences and death by suicide (although with no difference between EA-based quartiles), and we lacked information about the subjects’ *APOE* ε4 genotype.

In summary, our results indicate that BD is associated with a lower plasma Aβ_42/40_ ratio, specifically in those showing an accelerated EA, as well as a significant association between Aβ_42_ and EA acceleration in the postmortem prefrontal cortex of individuals with BD. This suggests that the observed variability in AD risk may be, at least to some extent, explained by EA acceleration in BD. Future studies should focus on individuals in the mid-to late-life age range and with larger sample sizes, exploring the clinical implications of these molecular alterations. Finally, this study provides preliminary evidence that targeting EA acceleration could ultimately reduce the risk of dementia and AD in individuals with BD.

## Data Availability

Datasets analyzed in this study and bioinformatic scripts will be made available from the corresponding author upon reasonable request.

## Author Contributions

GRF and JCS designed and conceptualized the study. JCS and GT were involved in sample recruitment and provided them for analyses. GRF, SDLG, NOZ, AWB, CNCL, and NK performed the experiments with all samples. GRF, SDLG, and NOZ performed all statistical analyses of the data. GRF wrote the first draft of the manuscript. TB, PES, BCD, and JCS edited the manuscript and provided input into data interpretation and analyses. All authors edited and approved the final version of the manuscript.

## Acknowledgments

We would like to thank the study participants for their willingness to participate in the study.

