## Supplementary Tables 2 - 4 and Figure S1 for "Association between epigenetic aging acceleration and amyloid biomarkers in bipolar disorder"

**Supplementary Table 2.** Comorbidities and medications in the N = 59 individuals with bipolar disorder.

| **Comorbidities** | **N (%)** |
| --- | --- |
| Generalized anxiety disorder | 5 (8.4%) |
| Post-traumatic stress disorder | 16 (27.1%) |
| Obsessive-compulsive disorder | 5 (8.4%) |
| Phobias | 21 (35.6%) |
| Personality disorder | 17 (28.8%) |
| Bulimia | 1 (1.7%) |
| Binge-eating disorder | 5 (8.4%) |
| Substance use disorder | 33 (55.9%) |
| **Medications** |  |
| Medication status, yes (%) | 51 (86.4%) |
| Lithium | 10 (16.9%) |
| Anticonvulsants | 27 (45.7%) |
| Antidepressants | 24 (40.6%) |
| Atypical antipsychotics | 31 (52.4%) |
| Typical antipsychotics | 3 (5.1%) |
| Benzodiazepines | 16 (27.1%) |
| Stimulants | 1 (1.7%) |

**Supplementary Table 3.** Associations between Alzheimer’s disease biomarkers and estimates of epigenetic aging acceleration in individuals with bipolar disorder and controls.

|  | **DunedinPACE** | | **AgeAccelGrim** | |
| --- | --- | --- | --- | --- |
|  | **R^2^** | **p-value** | **R^2^** | **p-value** |
| **Whole sample (N = 79)** | | | | |
| Aβ_40_ | 0.078 | 0.500 | 0.066 | 0.570 |
| Aβ_42_ | 0.120 | 0.270 | 0.058 | 0.610 |
| Total tau | -0.220 | 0.057 | -0.025 | 0.830 |
| Aβ_42/40_ | -0.200 | **0.008** | -0.140 | 0.071 |
| **Controls (N = 20)** | | | | |
| Aβ_40_ | -0.055 | 0.899 | 0.064 | 0.147 |
| Aβ_42_ | -0.015 | 0.410 | 0.057 | 0.161 |
| Total tau | 0.025 | 0.237 | -0.055 | 0.924 |
| Aβ_42/40_ | -0.010 | 0.379 | -0.035 | 0.557 |
| **Individuals with BD (N = 59)** | | | | |
| Aβ_40_ | -0.017 | 0.945 | -0.012 | 0.565 |
| Aβ_42_ | 0.006 | 0.254 | -0.016 | 0.803 |
| Total tau | 0.009 | 0.219 | -0.018 | 0.989 |
| Aβ_42/40_ | 0.005 | 0.259 | 0.001 | 0.306 |

**Supplementary Table 4.** Analysis of differences in quartiles of the DunedinPACE variable

| **Predictor** | **1^st^ quartile**  **(N = 15)** | **4^th^ quartile**  **(N = 15)** | **p-value** |
| --- | --- | --- | --- |
| Age, mean (SD) | 32.20 (8.89) | 34.73 (4.95) | 0.35^1^ |
| Sex, *n* Female (%) | 7 (47%) | 13 (87%) | 0.053^2^ |
| BMI, mean (SD) | 27.57 (6.01) | 34.95 (7.18) | **0.009**^1^ |
| Smoking score, median [IQR] | -0.68 [2.85] | 2.60 [8.11] | 0.2^3^ |
| Aβ_40_ (pg/mL), mean (SD) | 181.29 (30.18) | 178.50 (47.81) | 0.85^1^ |
| Aβ_42_ (pg/mL), mean (SD) | 10.05 (1.99) | 9.58 (2.17) | 0.54^1^ |
| Total Tau (pg/mL), mean (SD) | 3.94 (0.82) | 3.45 (1.54) | 0.28^1^ |
| Aβ_42/40_, median [IQR] | 0.055 [0.006] | 0.055 [0.006] | 0.72^3^ |
| AgeAccelGrim, median [IQR] | -0.56 [4.28] | 4.01 [4.54] | **0.008**^3^ |
| DunedinPACE, mean (SD) | 0.88 (0.09) | 1.25 (0.06) | **<0.001**^1^ |

Aβ – amyloid beta; BD – bipolar disorder; BMI – body mass index; IQR – interquartile range; SD – standard deviation. ^1^Welch Two Sample t-test; ^2^Pearson’s Chi-squared test; ^3^Wilcoxon rank sum exact test.

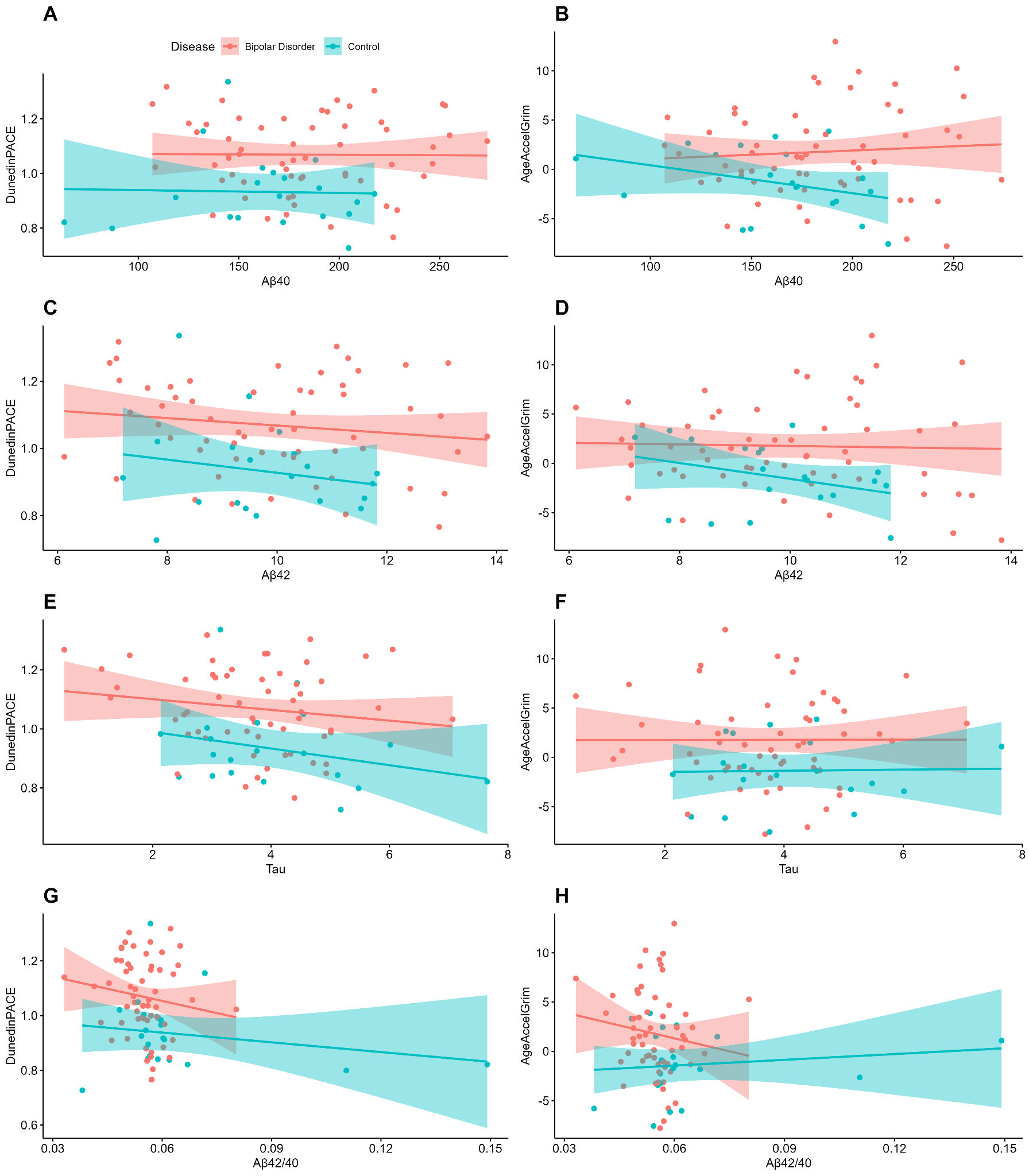

**Supplementary Figure 1.** Correlations between Alzheimer’s disease biomarkers and epigenetic aging acceleration estimates (AgeAccelGrim and DunedinPACE) in blood from individuals with bipolar disorder (red line, N = 59) and controls (blue line, N = 20). R-squared and p-values are shown in Supplementary Table 3.
